# The individual and combined impacts of school-based water, sanitation, and hygiene (WASH) education and infrastructure operation and maintenance (O&M): A factorial cluster-randomized controlled trial in Uttar Pradesh, India

**DOI:** 10.1101/2025.10.23.25338677

**Authors:** Gracie Hornsby, Christine Pu, Anvesh Badamikar, Poorva Shekher, Dinesh Bhatt, James Winter, Preksha Gupta, Nikita Purty, Ruhi Saith, Gary L. Darmstadt, Jennifer Davis

**Author notes:** co-Senior authors (Gary L. Darmstadt, Jennifer Davis). The Corresponding Author has the right to grant on behalf of all authors and does grant on behalf of all authors a worldwide licence to the Publishers and its licensees in perpetuity, in all forms, formats and media (whether known now or created in the future), to i) publish, reproduce, distribute, display and store the Contribution, ii) translate the Contribution into other languages, create adaptations, reprints, include within collections and create summaries, extracts and/or, abstracts of the Contribution, iii) create any other derivative work(s) based on the Contribution, iv) to exploit all subsidiary rights in the Contribution, v) the inclusion of electronic links from the Contribution to third party material where-ever it may be located; and, vi) licence any third party to do any or all of the above.* *The lead authors, Gary L. Darmstadt and Jennifer Davis, affirm that the manuscript is an honest, accurate, and transparent account of the study being reported; that no important aspects of the study have been omitted; and that any discrepancies from the study as planned have been explained.

## Abstract

**Introduction:** While improving water, sanitation, and hygiene (WASH) service delivery in schools supports the Sustainable Development Goals, rigorous trials have recently reported inconsistent effects of WASH in schools (WinS) programmes on student behaviour and health. We examined the individual and combined impacts of WASH education and infrastructure operation and maintenance (O&M) interventions on student knowledge, facility conditions, and hygiene behavior.

**Methods:** This four-arm cluster-randomized controlled trial (cRCT) was conducted over one year in 200 public primary schools of Uttar Pradesh, India. Pre-baseline infrastructure repairs were made to normalize access to WASH facilities in all schools. Interventions included a teacher-delivered, play-based WASH curriculum and a third-party O&M service including a part-time cleaner. Schools were randomized to Curriculum, O&M, Curriculum + O&M, or Control arms. We measured study outcomes with student and teacher surveys at three time points, and through unannounced behavior and infrastructure observations at nine and eight time points, respectively.

**Results:** The Curriculum intervention significantly improved all measured student WASH knowledge outcomes, whereas O&M alone showed no effect. Students in the Curriculum + O&M arm had the strongest gains, with 9.7 times greater odds of accurate germ knowledge (aOR: 9.67; p<0.001). The O&M intervention yielded significant and sustained improvements in toilet-related infrastructure outcomes, making toilets more likely to be unlocked, clean, functional, and to have water available for anal cleansing. Neither intervention, individually or combined, significantly changed observed rates of student toilet use or handwashing.

**Conclusion:** The curricular intervention was effective at improving students’ WASH-related knowledge, and the O&M intervention was effective at improving the quality of WASH service provision. These interventions, either alone or in combination, were insufficient to produce significant changes in student hygiene behavior. Future WinS research should consider the potential of normative behavior change theories.

**KEY MESSAGES:** *What is already known on this topic:*

● School-based interventions have produced mixed results on student behavior change, health, and absenteeism.
● Few prior studies have assessed the impact of improved O&M of school WASH facilities on service provision, student behavior, or health.

*What this study adds:*

● We found that student knowledge and toilet accessibility, functionality, and cleanliness improved; however, our study was novel in finding that neither WASH education nor WASH infrastructure O&M, alone or in combination, were sufficient to increase student handwashing and toilet use.
● These null results for student behavior change outcomes may help explain the numerous WinS studies that have found mixed or negligible changes in diarrhea, respiratory infection, and absenteeism.

*How this study might affect research, practice or policy:*

● Our results suggest that the researcher and practitioner community has not yet discovered how to reliably drive significant and scalable change in student WASH behaviors. Future research should aim to understand the mechanisms by which student behaviors change in the school environment to produce consistent and scalable benefits, including the role of norms.

## INTRODUCTION

The United Nations’ Sustainable Development Goals (SDGs) 4 and 6 call for equitable access to quality education and sustainable water and sanitation for all (1). Improving water, sanitation, and hygiene (WASH) service delivery in schools supports progress toward both goals. Schools are an important venue for providing WASH services because children spend a considerable amount of time in close contact with peers from other households. Schoolchildren are also particularly susceptible to infection with fecal pathogens that cause diarrhea (2) and respiratory infections (3) due to their immature immune systems and relative inexperience with good hygiene practices. In the school environment, these infections are associated with impaired school performance (4) and increased absenteeism (5), leading to downstream impacts on educational attainment (4,6,7) and future earnings (6,8).

For over three decades, governments in low- and middle-income countries (LMICs) have collaborated with development partners on ambitious WASH in schools (WinS) programs. While a 2019 review concluded that there is “no need for additional large-scale epidemiological studies on the impact of WinS on diarrhoea among students” given the strong evidence of positive impact (9), recent rigorous trials have found limited or mixed effects of WinS programs on diarrhea and respiratory illness in Addis Ababa (10) and Lao PDR (11). Some WinS studies have measured changes in students’ WASH behaviors instead of health outcomes, given the lower expense and ability to detect behavioral changes within shorter timeframes and smaller populations. The link between behaviors such as handwashing at critical times and reduction of open defecation with health have been well-established (3,12–16). Furthermore, measuring outcomes upstream from health provides an opportunity to test the underlying theory of change for an intervention.

Not only are the impacts of WinS on student health outcomes unclear; so too are the necessary and sufficient conditions for sustainable delivery of WASH services in schools. Many studies have investigated the impacts of school-based WASH education and new infrastructure on behavior (17–23), but far fewer have assessed the role of proactive operation and maintenance (O&M) of school WASH facilities (24–29). A recent systematic review reported that unclean or inoperable facilities stemming from poor O&M can deter student handwashing and latrine use, and highlighted the need for more evidence on effective O&M models (30). Critically, no published studies have tested the impacts of WASH educational and O&M interventions individually and combined, despite calls for more integrated WinS programs (31).

While several high-quality, large-scale studies have contributed valuable evidence on the impact of WinS investments, the broader literature is often constrained by small sample sizes (18–22,32), reliance on self-reported outcomes (18,20,21,23,27,33), and short measurement periods (18,20,22). To address these knowledge gaps, we aimed to rigorously quantify the individual and combined impacts of WASH education and enhanced infrastructure O&M on student knowledge, infrastructure conditions, and WASH-related behaviors.

## METHODS

### Study Design

We designed and implemented a prospective four-arm cluster-randomized controlled trial (cRCT) in Uttar Pradesh, India to measure along hypothesized causal pathways (Figure 1). The first pathway, inspired by the knowledge, attitudes, practices (KAPs) model for health behavior change (34), posits that delivery of a high-quality WASH curriculum will improve student knowledge of relevant WASH topics, which will in turn improve students’ WASH behaviors like handwashing and using a toilet to urinate and defecate. This pathway implies that students’ lack of knowledge is a key bottleneck to better WASH behaviors. The second pathway posits that implementation of an O&M intervention will improve the accessibility, cleanliness, and functionality of WASH infrastructure and the availability of consumables like water and soap. Improving the enabling environment will in turn improve students’ WASH behaviors.

**Figure 1.**
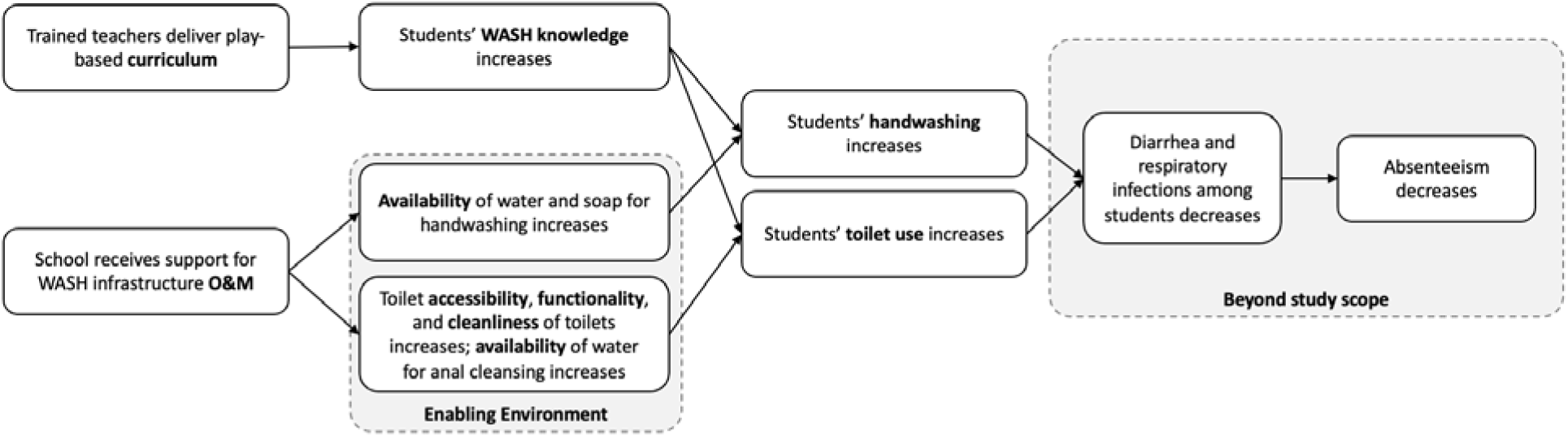
Conceptual model of how a WASH curriculum and O&M support lead to WASH-related behavior change in the school environment.

Similarly, this pathway implies that inaccessibility, uncleanliness, or non-functionality of WASH infrastructure or non-availability of water or soap are primary barriers to students washing hands and using the toilet. We also hypothesized that the combined impacts from both WASH education and an improved enabling environment will exceed their individual impacts. We posit that each pathway may be individually insufficient to markedly improve students’ behavior, but that both are necessary components of school-based WASH behavior change.

### Interventions

We evaluated two interventions. The first was a 12-session, play-based WASH curriculum targeting students in grades one through four. The curriculum was developed by a leading children’s education and media organization, and included six sessions of new content, followed by six sessions that reinforced the ideas presented in the first half of the program. Key learning objectives included identifying safe drinking water sources and critical times for handwashing, the importance of using a toilet, and practicing good personal hygiene. Teachers delivered the curriculum during regular school hours and received a package of teaching materials including games, play mats, storybooks, and videos with a miniature projector. After the baseline (pre-intervention) survey, 190 teachers completed a two-day, in-person training led by curriculum designers.

The second intervention was a WASH facility O&M service designed to improve cleanliness and functionality of school WASH facilities. Findings from formative research that informed the O&M intervention design will be published separately (35). In brief, each school was assigned one part-time cleaner called a *Swachhata Saathi* (Cleanliness Buddy) who received an initial two-day training and was responsible for cleaning in and around the school’s water source, handwashing station(s), and toilets. The *Swachhata Saathi* was also charged with replenishing soap and water at handwashing stations, ensuring that toilets had water available for anal cleansing, and reporting non-functional infrastructure or out-of-stock soap to school staff. Headmasters agreed that their school would be responsible for providing soap for student handwashing. Additional intervention details are in Supplementary Section S2.

### Outcomes

Our pre-specified primary outcomes were the observed rates of student toilet use and student handwashing with water and/or soap, defined as the count of students observed practicing the target behavior during a randomly selected two-hour observation period divided by the estimated daily attendance of the school. These outcomes were measured during eight unannounced observations per school.

We also measured nine pre-specified intermediate outcomes: student knowledge of two critical handwashing times and germs, toilet usability (unlocked, cleanliness score, functionality), and availability of water for anal cleansing and soap and water for handwashing. We established definitions and measurement rules for intermediate outcomes (Table S1) based on prior WinS studies reported in Pu et al. to promote comparability across trials (36).

### Sampling Frame

The study site included five non-contiguous community development blocks (hereafter, ‘block’) of one district of Uttar Pradesh that will not be identified to protect participants’ privacy. Given our research objectives and study design, a sample size of 50 schools per arm (200 total) was sufficient to detect effect sizes of 0.07 on measures of infrastructure functionality and 0.17 on student-level outcomes with 95% confidence and 80% power. Figure 2 summarizes our school selection, randomization, and survey sampling process. Additional information on power calculations and sampling is provided as Supplementary Information (Sections S3 and S4).

**Figure 2.**
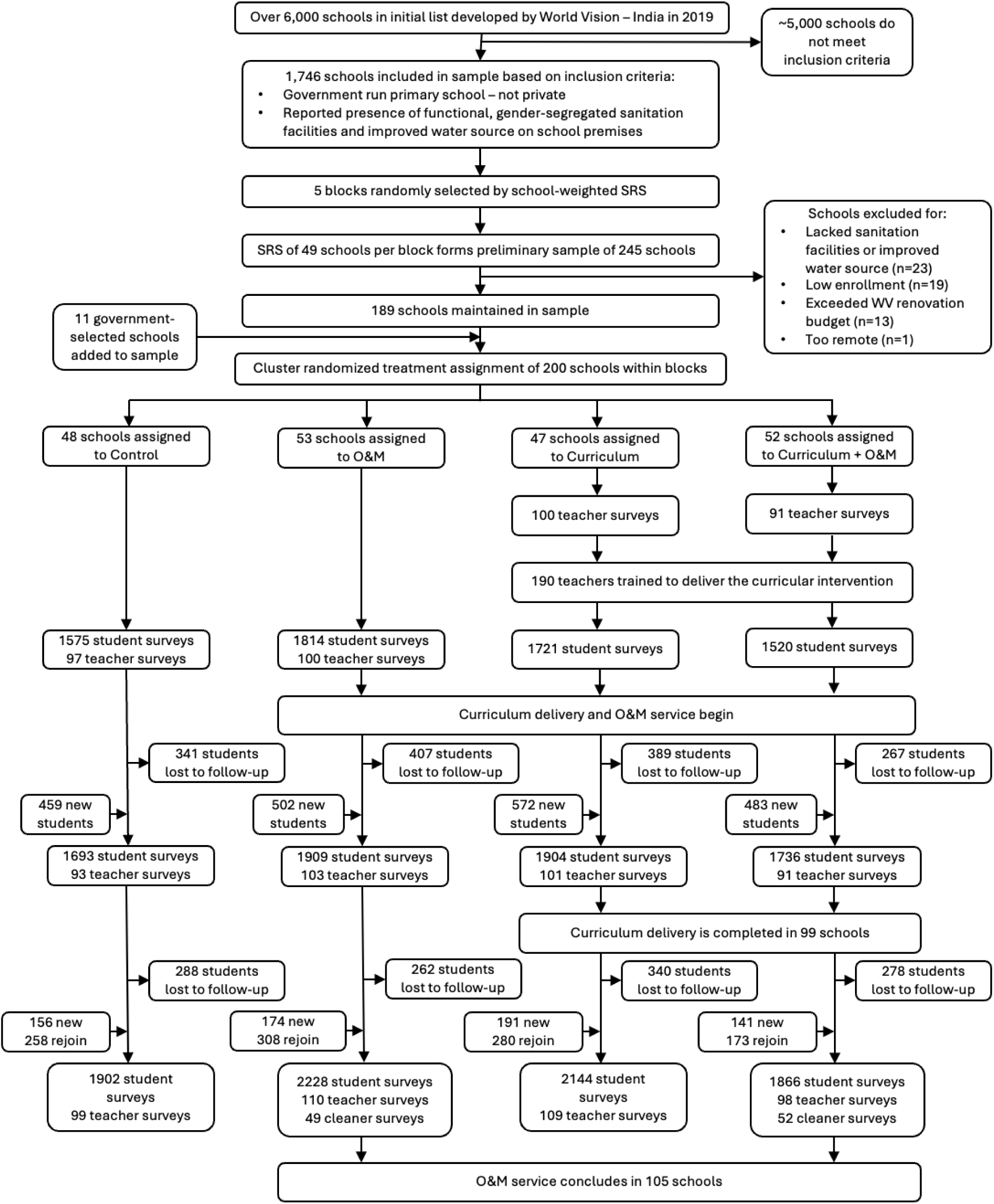
Trial profile. There were 23 schools that did not meet infrastructure eligibility criteria for the study but were not screened out from the parent population of over 6,000 schools using information from the national Unified District Information System for Education (UDISE) dataset. When subsequent in-person visits revealed errors in the UDISE dataset, these schools were removed from the preliminary sample of 245 schools.

Contracted technicians undertook repairs or upgrades to school WASH infrastructure as needed to ensure that every study school had an on-premises water source, handwashing station, and gender-segregated toilet facility prior to the start of the study to create comparable baseline access to WASH facilities.

### Treatment Assignment and Blinding

We used cluster randomization to control for unmeasured confounders across blocks, keep implementation logistics feasible, and reduce the risk of spillover between treatment and nearby control schools. Using a random number generator, we assigned each cluster to one of the four study arms: (1) WASH curriculum plus O&M (“Curriculum + O&M”), (2) WASH curriculum only (“Curriculum”), (3) O&M only (“O&M”), and (4) Control. Because clusters had different numbers of schools, our sampling strategy resulted in 52, 47, 53, and 48 schools being assigned to arms 1 through 4, respectively (Figure 2). Additional information on treatment assignment is provided in Supplemental Section S5.

School staff and field staff could not be blinded to the treatment assignment of each school due to the visible nature of both interventions. All members of the research team remained blinded to treatment assignment until all pre-specified analyses were complete.

#### Data Collection

Both interventions were launched during the autumn of 2022, immediately after all baseline measurements were collected. We conducted midline measurements in all schools roughly six months later, after all key WASH concepts should have been introduced in schools receiving the WASH curriculum intervention. We took endline measurements roughly one year after baseline when schools should have completed all 12 sessions of the curriculum. The O&M intervention was delivered until all endline measurements were completed.

We collected data from students, teachers, and cleaners at each school using structured surveys in CSPro (version 7.7). We targeted students and teachers in grades one and four because they are the highest and lowest grades for which the curriculum was designed. At each school, we aimed to survey two teachers directly involved in delivering the WASH curriculum, 20 grade one students, and 20 grade four students during each round of data collection. Additional information on student selection is provided in Supplemental Section S6.

We made unannounced observations of school WASH infrastructure functionality and cleanliness at baseline, midline, and endline, as well as every 2-3 weeks between rounds of surveying (n=11 observations per school in total). We also completed unannounced, two-hour observations of student toilet use and handwashing behavior at baseline, midline, and endline, on a different day than student and teacher surveying since surveying likely disturbed students’ normal schedule. Additional student behavior observations were completed every 3-4 weeks between survey rounds (n=10 per school in total). Infrastructure and behavior observations were recorded with pencil and paper and entered into CSPro on field tablets after leaving the school premises.

### Statistical Analyses

We registered a pre-analysis plan with 3iE’s Registry for International Development Impact Evaluations (RIDIE) prior to starting data analysis. We described and submitted deviations from this original analysis plan to RIDIE for prior to publication (RIDIE-STUDY-ID-662bed52ae1e7).

We conducted all analyses using an intention-to-treat approach with a difference-in-differences (DiD) framework for all models. We used linear mixed models based on the distribution of each outcome to model knowledge, infrastructure, and behavior. A summary of our analytical approach is provided in Table S4. Additional information on data analysis, including statistical adjustment of baseline data, is provided in Supplementary Sections S7 and S8. We accounted for the hierarchical nature of these data with random effects and computed cluster-robust standard errors to produce more conservative p-values for all models. Moreover, we applied a multiple testing adjustment for all p-values using the Benjamini-Hochberg approach to reduce the chance of a Type 1 error (37). We carried out all analyses in R Version 2023.06.0+421 and stored all data in a secure online repository.

### Ethics & Role of the Funding Source

We obtained ethical approval from Stanford University (California, USA, protocol 69912) and the Sigma IRB in India (protocol 10046/IRB/22-23). The research was explained to, and permission to engage with schools was obtained from, Uttar Pradesh Department of Education officials. We obtained written consent from headmasters, teachers, and each student’s parent at the beginning of the study. Teachers explained consent forms to parents. In instances where parents were not literate, we collected fingerprints as signatures. We obtained oral assent from students prior to every survey. Control schools received the curricular intervention after endline measurements concluded.

World Vision USA (WV) funded the study, including the initial repairs and upgrades to school WASH infrastructure. They had input into conceptualizing the study, articulating a causal model, formulating the research questions and choosing the study site. WV did not participate in data collection, analysis, or interpretation, nor in writing the manuscript.

### Patient and public involvement

Research participants and the public were not involved in study design. School headmasters and teachers provided input about the hiring of *Swachhata Saathis.* Data collection instruments were piloted with teachers and students outside the study sample and revised based in part on their feedback. Study findings were disseminated to governmental and school staff and parents through community meetings.

## RESULTS

All 200 schools in the trial assigned to the Curriculum, O&M, and Curriculum + O&M treatments consented to participate and undergo randomization to receive treatment. No school withdrew from the study entirely, although one Control school refused to continue with infrastructure and behavior observations after two rounds of observation. As a result, only the student and teacher survey data from this school were included in our analyses. All (100%) Curriculum and Curriculum + O&M schools reported completing all 12 sessions of the curricular intervention. A cleaner was successfully hired and employed for the entire study duration at all 105 O&M and Curriculum + O&M schools.

Following baseline data collection, we performed balance checks to assess if our cluster randomization created comparable study arms. We found significant differences in observed baseline soap availability (Tables S3 and S5). However, this outcome was omitted from analysis since data were too sparse to model. Other school-and student-level variables were statistically similar across treatment groups at baseline (Table S5).

A total of 6,630, 7,242, and 8,140 students were surveyed at baseline, midline, and endline, respectively. Fifty-eight percent (n=4,761) of the student sample participated in all three rounds of surveying, 39% (n=3,182) participated in two rounds of surveying, and 17% (n=1,365) completed only one survey. Of the students enrolled in the study, 49% were girls and 51% were boys per our gender-stratified sampling strategy; 48% of the baseline students were in grade one and 52% in grade four. Two-thirds (66%) of students in our sample reported having access to a toilet at home and 84% reported “almost always” having access to soap for handwashing at home.

### Student Knowledge Outcomes

Schools that received the curricular intervention made significant improvement on all measured student WASH knowledge outcomes (Table 1). By contrast, little or no gains in student knowledge were observed in schools that received the O&M intervention only. Students in all three intervention arms had significantly increased odds of identifying “before eating” as a critical handwashing time relative to students in the Control arm, despite high baseline knowledge (predicted probability of 0.90-0.94). At endline, students in the Curriculum arm were nearly twice as likely as those in the Control arm to identify this time in response to an open-ended question (adjusted odds ratio [aOR] = 1.89; 95% CI: 1.27–2.81; p = 0.004). The Curriculum + O&M arm showed the largest improvement with students being more than twice as likely to identify handwashing before eating as a critical time (aOR = 2.74; 95% CI: 1.81–4.16; p < 0.001). The O&M arm also demonstrated significantly higher odds (aOR = 1.89; 95% CI: 1.29–2.78; p = 0.003), though this translated to only a 2 percentage point increase in probability. The statistical significance of this result may be attributable to its high baseline value (0.94), as relatively small changes in probability in the tail of the log-odds distribution can lead to large changes in log-odds.

**Table 1.** Intervention effects on primary and secondary outcomes. All model results were calculated in terms of change in log-odds. For easier interpretation, we converted these estimates to adjusted odds ratios (aOR) using the contrast() function in R. To better illustrate time-varying results, findings are also presented as predicted probabilities, calculated using the emmeans package in R. Since student knowledge outcomes displayed roughly linear trends, we report endline effects to summarize change over the duration of the trial. For infrastructure and behavior outcomes which displayed more fluctuation and non-linearity, we calculated the group-time average treatment effect (ATE) using asymptotic theory (37) to estimate the average effect of treatment across the entire intervention period.

| Outcome | Control | Curriculum |  |  | O&M |  |  | Curriculum + O&M |  |  |
| --- | --- | --- | --- | --- | --- | --- | --- | --- | --- | --- |
|  | BL Prob. (SE) | DiD change | aOR (95% CI) | p-value | DiD change | aOR (95% CI) | p-value | DiD change | aOR (95% CI) | p-value |
| HW before eating | 0.94 (0.01) | +0.07 | 1.89 (1.27, 2.81) | 0.004 * | +0.02 | 1.89 (1.29, 2.78) | 0.003 * | +0.05 | 2.74 (1.81, 4.16) | 0.000 * |
| HW after toilet use | 0.57 (0.04) | +0.04 | 1.78 (1.12, 2.82) | 0.030 * | -0.01 | 1.35 (0.87, 2.11) | 0.291 | +0.08 | 2.13 (1.35, 3.34) | 0.003 * |
| Germ knowledge | 0.08 (0.02) | +0.39 | 7.54 (3.13, 18.16) | 0.000 * | +0.02 | 1.37 (0.60, 3.13) | 0.519 | +0.45 | 9.67 (4.10, 22.83) | 0.000 * |
|  | BL Prob. (SE) | DiD change | aOR (95% CI) | p-value | DiD change | aOR (95% CI) | p-value | DiD change | aOR (95% CI) | p-value |
| Toilets unlocked | 0.73 (0.06) | +0.00 | 0.93 (0.60, 1.43) | 0.770 | +0.20 | 2.72 (1.72, 4.31) | 0.000 * | +0.20 | 3.72 (2.28, 6.07) | 0.000 * |
| Toilet flushes | 0.51 (0.08) | -0.10 | 0.72 (0.44, 1.18) | 0.291 | +0.30 | Inf (Inf, Inf)<sup>a</sup> | 0.000 * | +0.10 | Inf (Inf, Inf)<sup>a</sup> | 0.000 * |
| Anal cleansing water available | 0.32 (0.07) | +0.00 | 1.12 (0.70, 1.80) | 0.295 | +0.20 | 2.99 (1.95, 4.59) | 0.000 * | +0.20 | 2.49 (1.60, 3.88) | 0.000 * |
|  | BL Score (SD) | DiD change | aOR (95% CI) | p-value | DiD change | aOR (95% CI) | p-value | DiD change | aOR (95% CI) | p-value |
| Toilet cleanliness score | 4.1 (2.4) | -0.5 | 0.91 (0.74, 1.11) | 0.436 | +3.5 | 2.09 (1.69, 2.58) | 0.000 * | +2.6 | 1.73 (1.40, 2.12) | 0.000 * |
|  | BL Prob. (SE) | DiD change | aOR (95% CI) | p-value | DiD change | aOR (95% CI) | p-value | DiD change | aOR (95% CI) | p-value |
| Observed student toilet use | 0.02 (0.01) | -0.010 | 1.20 (0.80, 1.80) | 0.448 | +0.013 | 1.84 (1.07, 3.16) | 0.055 | +0.016 | 1.68 (1.04, 2.71) | 0.065 |
| Observed student handwashing | 0.10 (0.02) | +0.010 | 0.97 (0.76, 1.24) | 0.809 | +0.007 | 1.03 (0.80, 1.33) | 0.809 | +0.006 | 1.09 (0.83, 1.43) | 0.590 |

\* Denotes a significant p-value (p<0.05) after the Benjamini-Hochberg adjustment. P-values are associated with aOR's.
<sup>†</sup> The aOR is estimated as 'Inf' since 100% of schools had flushable (functional) toilets at some measurements.
Note: The toilet cleanliness score ranges from 0-8
Note: The aOR is the exponentiated form of the log-odds, representing the proportional change in the odds of an outcome for the treatment arm as compared to the Control.
Note: Predicted probabilities range continuously from 0-1 and represent the estimated likelihood of the outcome on a linear scale.

|  | Control | Curriculum |  |  | O&M |  |  | Curriculum + O&M |  |  |
| --- | --- | --- | --- | --- | --- | --- | --- | --- | --- | --- |
| Outcome | BL Prob. (SE) | DiD change | aOR (95% CI) | p-value | DiD change | aOR (95% CI) | p-value | DiD change | aOR (95% CI) | p-value |
| HW before eating | 0.94 (0.01) | +0.07 | 1.89 (1.27, 2.81) | 0.004 * | +0.02 | 1.89 (1.29, 2.78) | 0.003 * | +0.05 | 2.74 (1.81, 4.16) | 0.000 * |
| HW after toilet use | 0.57 (0.04) | +0.04 | 1.78 (1.12, 2.82) | 0.030 * | -0.01 | 1.35 (0.87, 2.11) | 0.291 | +0.08 | 2.13 (1.35, 3.34) | 0.003 * |
| Germ knowledge | 0.08 (0.02) | +0.39 | 7.54 (3.13, 18.16) | 0.000 * | +0.02 | 1.37 (0.60, 3.13) | 0.519 | +0.45 | 9.67 (4.10, 22.83) | 0.000 * |
|  | BL Prob. (SE) | DiD change | aOR (95% CI) | p-value | DiD change | aOR (95% CI) | p-value | DiD change | aOR (95% CI) | p-value |
| Toilets unlocked | 0.73 (0.06) | +−0.00 | 0.93 (0.60, 1.43) | 0.770 | +0.20 | 2.72 (1.72, 4.31) | 0.000 * | +0.20 | 3.72 (2.28, 6.07) | 0.000 * |
| Toilet flushes | 0.51 (0.08) | -0.10 | 0.72 (0.44, 1.18) | 0.291 | +0.30 | Inf (Inf, Inf)<sup>a</sup> | 0.000 * | +0.10 | Inf (Inf, Inf)<sup>a</sup> | 0.000 * |
| Anal cleansing water available | 0.32 (0.07) | +0.00 | 1.12 (0.70, 1.80) | 0.295 | +0.20 | 2.99 (1.95, 4.59) | 0.000 * | +0.20 | 2.49 (1.60, 3.88) | 0.000 * |
|  | BL Score (SD) | DiD change | aOR (95% CI) | p-value | DiD change | aOR (95% CI) | p-value | DiD change | aOR (95% CI) | p-value |
| Toilet cleanliness score | 4.1 (2.4) | -0.5 | 0.91 (0.74, 1.11) | 0.436 | +3.5 | 2.09 (1.69, 2.58) | 0.000 * | +2.6 | 1.73 (1.40, 2.12) | 0.000 * |
|  | BL Prob. (SE) | DiD change | aOR (95% CI) | p-value | DiD change | aOR (95% CI) | p-value | DiD change | aOR (95% CI) | p-value |
| Observed student toilet use | 0.02 (0.01) | -0.010 | 1.20 (0.80, 1.80) | 0.448 | +0.013 | 1.84 (1.07, 3.16) | 0.055 | +0.016 | 1.68 (1.04, 2.71) | 0.065 |
| Observed student handwashing | 0.10 (0.02) | +0.010 | 0.97 (0.76, 1.24) | 0.809 | +0.007 | 1.03 (0.80, 1.33) | 0.809 | +0.006 | 1.09 (0.83, 1.43) | 0.590 |

\* Denotes a significant p-value (p<0.05) after the Benjamini-Hochberg adjustment. P-values are associated with aOR's.
† The aOR is estimated as 'Inf' since 100% of schools had flushable (functional) toilets at some measurements.
Note: The toilet cleanliness score ranges from 0-8
Note: The aOR is the exponentiated form of the log-odds, representing the proportional change in the odds of an outcome for the treatment arm as compared to the Control.
Note: Predicted probabilities range continuously from 0-1 and represent the estimated likelihood of the outcome on a linear scale.

Students’ knowledge of the importance of handwashing after toilet use was lower at baseline (predicted probability of 0.57-0.63) than for before eating. At endline, students in the Curriculum arm had 1.78 times greater odds of identifying “after using the toilet” as a critical handwashing time compared to the Control (aOR = 1.78; 95% CI: 1.12–2.82; p = 0.030), while the Curriculum + O&M arm showed even higher odds (aOR = 2.13; 95% CI: 1.35–3.34; p = 0.003). These corresponded to 4 and 8 percentage point increases, respectively.

Students’ baseline knowledge of germs and their relevance to WASH and health was low across all arms (predicted probability of 0.18-0.25) and increased markedly in arms that received the curriculum. Curriculum arm students had 7.5 times greater odds of providing accurate and relevant germ knowledge at endline than those in the Control group (aOR = 7.54; 95% CI: 3.13–18.16; p < 0.001), corresponding to a 39 percentage point increase in probability. The Curriculum + O&M arm showed an even stronger effect (aOR = 9.67; 95% CI: 4.10–22.83; p < 0.001), equivalent to a 45 percentage point increase in probability.

We detected no differences at any time points in the share of boys versus girls who reported “before eating” and “after using the toilet” as critical handwashing times (Figures S2 and S3). Girls reported significantly lower baseline germ knowledge than boys (predicted probability of 0.14 versus 0.18, p<0.001); however, both groups’ knowledge of germs improved at similar rates thereafter (Figure S4). Similarly, grade one students had significantly lower baseline germ knowledge than grade four students. While we detected a significant grade-by-time effect in the model, the magnitude of change in predicted probability was the same for grade one and grade four students, indicating that both grades benefitted similarly from the program.

### School Infrastructure and Consumables Outcomes

The O&M intervention had a significant effect on all toilet-related WASH infrastructure and consumables outcomes (Figure 3) and these impacts were generally sustained throughout the study (Figure S5). By contrast, schools that received only the curricular intervention demonstrated no significant impacts. Data from unannounced spot checks indicate that toilets in the O&M arm had 2.72 times higher odds of being unlocked across all post-baseline measurements (avg. aOR = 2.72; 95% CI: 1.72–4.31; p < 0.001), while the Curriculum + O&M arm showed an even larger effect (avg. aOR = 3.72; 95% CI: 2.28–6.07; p < 0.001).

**Figure 3.**
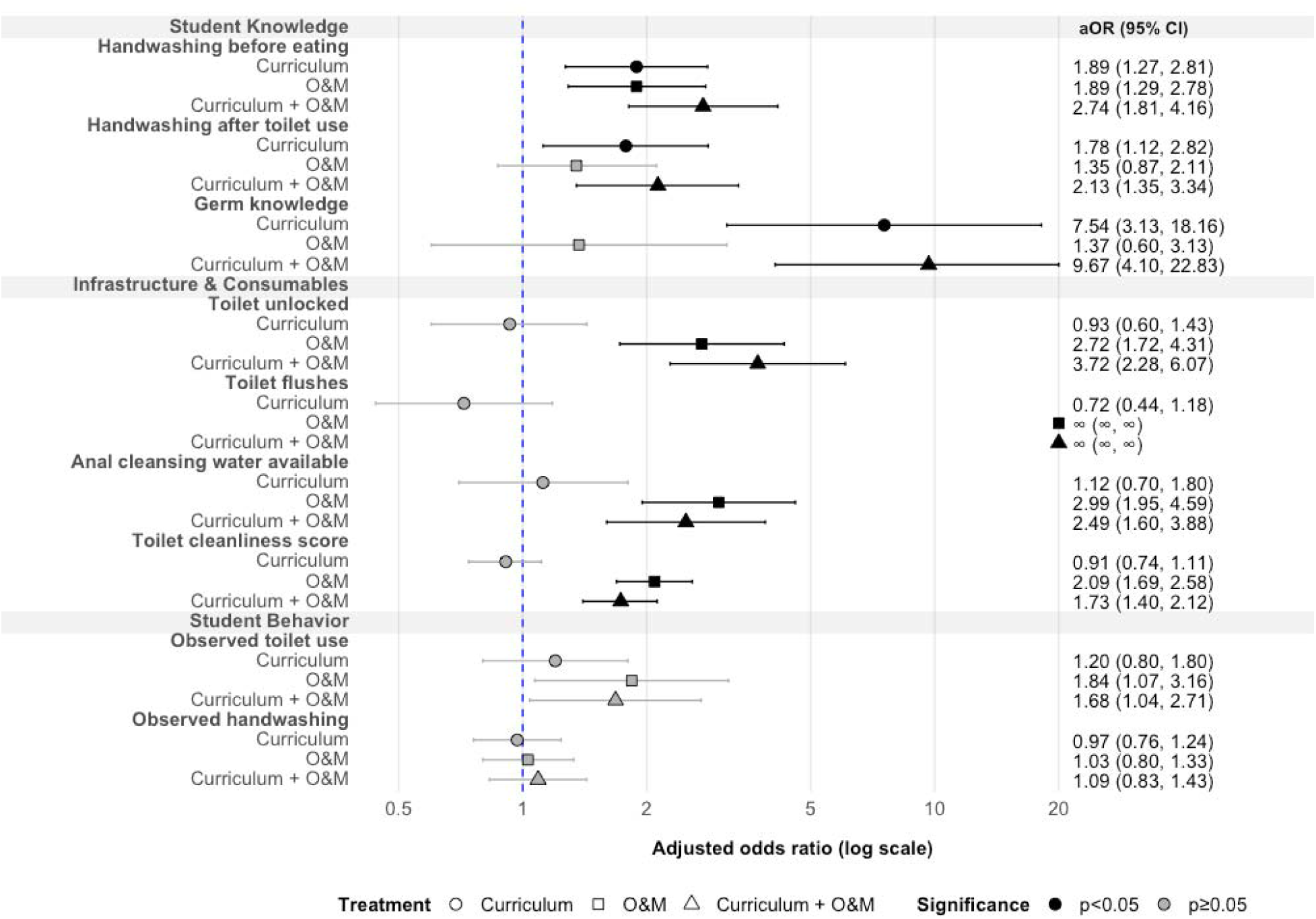
Forest plot displaying results of primary and second outcomes. All results are displayed as adjusted odds ratios. The x-axis is on a logarithmic scale to facilitate the visualization of a wide range of effect sizes. Some aORs are estimated as infinity since 100% of schools had flushable (functional) toilets at some measurements. Confidence intervals for O&M and Curriculum + O&M arms for observed student toilet use do not cross zero but are not colored as statistically significant since the p-value was not statistically significant after adjustment for multiple testing.

Toilets in the O&M and Curriculum + O&M arms also had significantly (p<0.001) higher odds of flushing properly. Toilet cleanliness scores (scored 0-8; definition section S9) also improved in the O&M arm by 3.5 points (avg. aOR = 2.09; 95% CI: 1.69–2.58; p < 0.001), while the score in the Curriculum + O&M arm improved by an average of 2.6 points (avg. aOR = 1.73; 95% CI: 1.40–2.12; p < 0.001).

Water for anal cleansing was also more frequently available in the O&M arms. The odds of toilets having adequate water were approximately 3 times higher in the O&M arm (avg. aOR = 2.99; 95% CI: 1.95–4.59; p < 0.001) and 2.5 times higher in the Curriculum + O&M arm (avg. aOR = 2.49; 95% CI: 1.60–3.88; p < 0.001). Although the O&M service was not designed to directly address water point functionality or soap availability, we still measured these outcomes because the *Swachhata Saathis’* cleaning and reporting duties may have indirectly influenced them, and because they in turn influence students’ ability to wash their hands. We found that, despite headmasters agreeing to provide soap for student handwashing, soap was rarely available in any study arm (8-29% of schools at each unannounced spot check), which resulted in availability data being too sparse to model. By contrast, 95-99% of schools had adequate water for handwashing at each time point, which also resulted in insufficient heterogeneity to model.

### Student Behavior Outcomes

The Curriculum, O&M, and Curriculum + O&M interventions had no significant impacts on rates of observed student toilet use or handwashing with water and/or soap (Figure 4).

**Figure 4.**
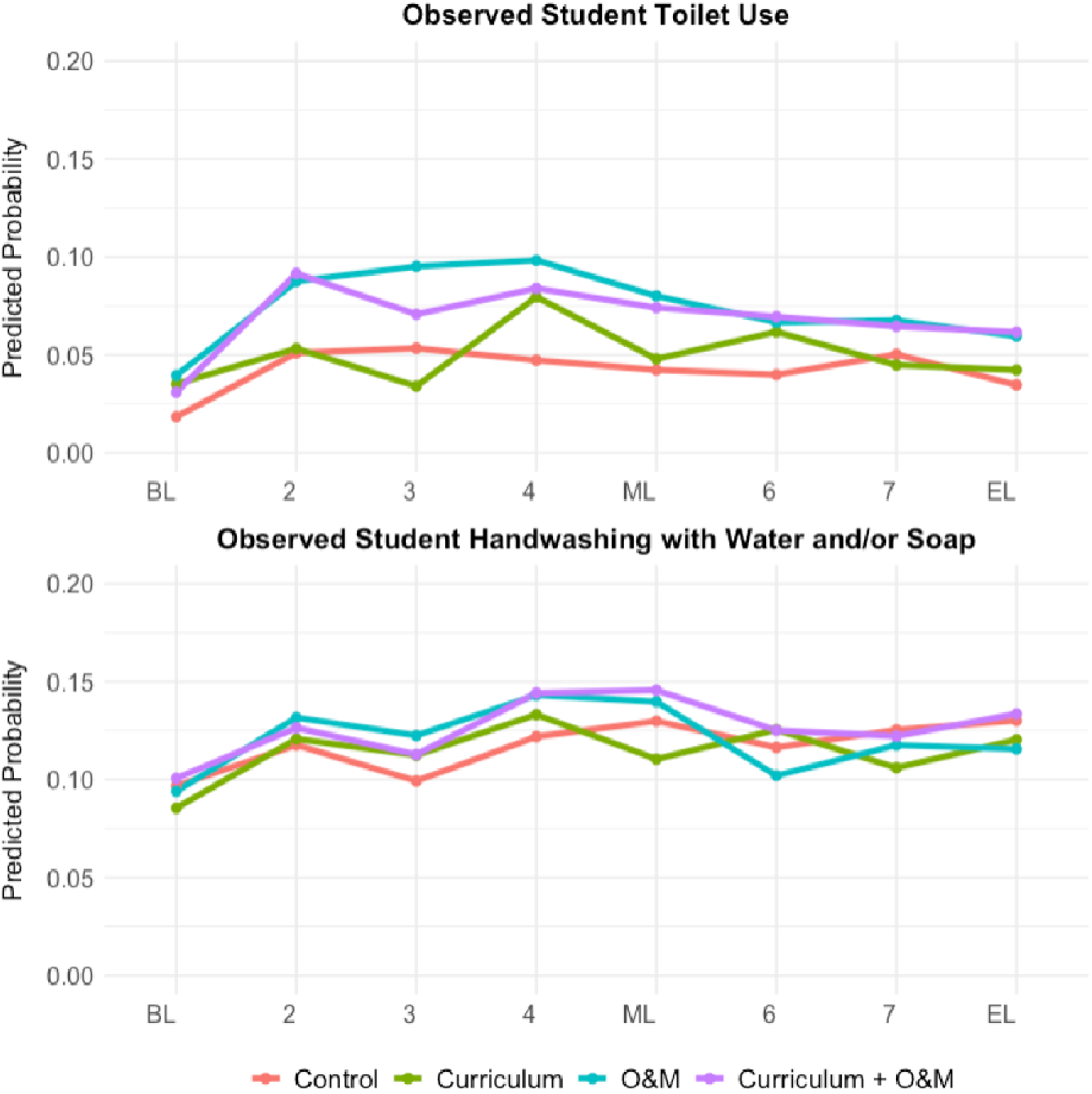
Time-varying results in units of predicted probability for primary outcomes, observed student toilet use and observed student handwashing with water and/or soap. “BL,” “ML,” and “EL” refer to baseline, midline, and endline (unannounced) observations which were performed on a different day than student and teacher surveys. Numbered measurements refer to unannounced behavior observations performed between major rounds of surveying.

Relative to the Control, the odds of an additional student using the school toilet tended to be higher in the O&M (avg. aOR 1.84, CI 1.07-3.16, p=0.055) and Curriculum + O&M arms (avg. aOR 1.68, CI 1.04-2.71, p=0.065); however, these estimates corresponded to only a 1–2 percentage point increase in the probability of a student using the school toilet and did not remain significant after adjusting for multiple testing. The Curriculum arm showed no significant effect (avg. aOR 1.20, CI 0.81-1.80, p=0.448). All three intervention arms performed similarly to the Control group on observed student handwashing with water and/or soap (Curriculum avg. aOR 0.97, CI 0.76-1.24, p=0.809; O&M avg. aOR 1.03, CI 0.80-1.33, p=0.809; Curriculum + O&M avg. aOR 1.09, CI 0.83-1.43, p=0.590) (Figure 3). Sensitivity analyses confirmed the robustness of our findings (Supplemental Section 10 and Table S6).

## DISCUSSION

Our factorial, cluster randomized trial in 200 public schools in India was uniquely designed to test the individual and combined effects of WASH education and infrastructure O&M interventions on knowledge, infrastructure, and behavior. We found that the WASH curriculum, both alone and in combination with the infrastructure O&M service, significantly improved students’ WASH knowledge; and the infrastructure O&M service, both alone and in combination with the curricular intervention, improved the quality of WASH service provision. However, both interventions, alone and in combination, failed to generate measurable change in student behavior.

The clearest evidence of intervention effectiveness was the improvement in students’ knowledge of critical handwashing times and germ theory. Specifically, students in the Curriculum group were twice as likely to identify handwashing after toilet use and before eating as critical times. Furthermore, they were 7-to 10-times more likely than students in Control schools to demonstrate accurate germ knowledge in an open-ended question. We believe that this understanding of germ theory indicates that students are developing the foundational knowledge to reason about future health and hygiene decisions beyond the explicit content of the curriculum. These findings were consistent for boys and girls, which is notable since the curriculum featured an empowered female lead character teaching a male character. Our findings were also consistent across grades one and four, indicating the curriculum’s effectiveness across its target age range. Notably, knowledge gains were consistently higher when the education and O&M interventions were implemented concurrently. We hypothesize that improved content retention may be a result of its greater salience in a school environment which reflects the hygienic conditions described in the curriculum. However, this requires further quantitative and qualitative investigation.

Our findings are consistent with many studies which report that school-based WASH education is effective at improving student knowledge (20,21,38–43). However, in contrast with prior literature (17,19), we found no evidence that the WASH curriculum alone improved school facilities, nor that it enhanced the effectiveness of O&M on WASH infrastructure outcomes when the two were implemented in tandem.

Our data also show that the O&M intervention resulted in rapid, significant and sustained improvements to school WASH infrastructure. Toilets in O&M schools were two to four times more likely to be unlocked and have water for anal cleansing, on average two to four points cleaner (out of eight), and far more likely to be functional than those in Control schools. Since the *Swachhata Saathis* were not equipped to perform repairs, this suggests that enhanced cleaning may have acted as a form of preventative maintenance, averting breakdown and promoting uninterrupted service.

Our O&M service was designed to provide continuous, proactive support with resources, bi-directional information flows, and meaningful accountability mechanisms (35). In contrast, the majority of O&M interventions tested in the literature included one-off trainings or disbursements of funds and consumables, with more variable results (24–26,26–28,44–46). Additional research is needed on effective public and private O&M arrangements that address the resource, information, and accountability (36) gaps that schools experience. In particular, arrangements that can be integrated into the existing school funding and management system may be more sustainable and scalable.

Despite improvements in students’ WASH-related knowledge and school WASH infrastructure conditions, we detected no significant effects on the rate of students’ toilet use or handwashing. Average rates of toilet use (predicted probability 0.01-0.04) and handwashing (predicted probability 0.08-0.10) were relatively low at baseline and remained low throughout the study period. This finding was consistent across both individual interventions and the combined treatment arm.

The lack of impact on student WASH behaviors is particularly interesting given that it refutes the original hypothesis of this trial. We anticipated that the individual interventions might be insufficient to spark change, but hypothesized that improving both knowledge and the enabling environment with a combination of curriculum and O&M support could spur meaningful shifts in behavior. Our results thus suggest that our original causal model may be flawed in its assumptions about if and how knowledge improvements and enhanced O&M translate into WASH-related behavior change. Other high-quality studies have similarly found it challenging to change students’ handwashing and toilet use habits with these types of interventions (24,47–50), suggesting that the path from educational programming and O&M to student action is more complex than previously understood. This mounting body of evidence on student behavior change may also help explain the numerous studies that have sought to measure changes in diarrhea, respiratory infection, and absenteeism due to WinS programming but have produced mixed or null results (10,24,51–53).

An important caveat regarding our behavior results concerns the lack of variation in the enabling environment for handwashing across study arms. Water was almost universally available but soap was rarely observed across all schools, despite headmasters reporting no significant barriers to providing soap during formative work and committing to provide soap for student handwashing as a condition of receiving the O&M intervention. We were thus prevented from testing the individual and combined impacts of handwashing materials availability on student handwashing, as depicted in Figure 1. Therefore, our handwashing results primarily reflect the impact of our other causal pathway, the individual impact of the curricular intervention.

The rare soap availability in this trial is particularly noteworthy since the curricular intervention specifically emphasized the importance of handwashing with soap. We could speculate that students might have been more likely to translate their knowledge to action if they were able to practice handwashing with soap rather than only water. However, additional analysis showed that handwashing rates were not systematically higher in schools where soap was observed than in schools where only water was available (unpublished data). Other studies have tested the impact of implementing hand hygiene education in the presence of adequate soap and water in schools in Bangladesh (54,55), the Philippines (47,56), and Kenya (50) and reported mixed results.

### Strengths and Limitations

Strengths of this study include the large sample size of both schools and students, long study duration, pre-specified causal pathways and outcomes in a registered pre-analysis plan, as well as the use of structured observations rather than reported data to monitor infrastructure and behavior outcomes.

There were also several important limitations. We chose to target student’s independent hygiene choices rather than group hygiene activities, like group handwashing before a meal, which is common in the context of India’s mid-day meal program. Thus, our observed behavioral data do not capture potential increases in handwashing during teacher-facilitated, high-traffic periods. Observations of toilet functionality, cleanliness, and availability of water for anal cleansing were limited to toilets that were unlocked upon our arrival, leaving us without data on these attributes for locked toilets. The observed soap data also had insufficient heterogeneity to be modeled as an intermediate outcome of the treatment. Consequently, we had to change our primary handwashing outcome from handwashing with soap to handwashing with water and/or soap, even though handwashing with water alone is less effective at removing fecal pathogens (57). Other limitations include our block selection method, which was based on the number of schools rather than students and biased our sample toward smaller schools, as well as the inclusion of 11 non-randomly selected schools, although sensitivity analysis suggests this had no effect on our results.

## CONCLUSIONS

Whereas the curricular intervention was highly effective at improving students’ WASH-related knowledge, our results suggest that knowledge formation alone was insufficient to produce significant changes in health behavior in this context. Further research is warranted to understand the long-term effects of early-childhood WASH education on behavior, health, and educational attainment. Similarly, the O&M intervention was effective at improving the accessibility, functionality, and cleanliness of toilets, as well as water availability for anal cleansing. Functional and clean WASH infrastructure is a crucial precondition for habit formation among students since certain facilities have to be present for students to practice healthy WASH behaviors. However, enhanced O&M alone proved insufficient to improve behavior in this trial.

Most importantly, and a unique strength of this study, we found that delivering these interventions in tandem was still insufficient to produce significant changes in student toilet use and handwashing at school. These interventions were well-resourced, high-quality, well-implemented, and they had significant impacts across all intermediate outcomes we expected them to affect. We assert that these interventions were not ineffective; rather, our findings alongside other recent WinS literature suggest that the research and practitioner community has not yet discovered how to reliably drive significant and scalable change in student WASH behaviors. We should not expect these programs to consistently yield meaningful impacts on student behavior, health, and absenteeism until we better understand these mechanisms of change.

Behavioral change is a multi-faceted and multi-factorial process, and the WASH community can take insights from other fields such as social psychology and organizational behavior to explore other pathways to student behavior change. For example, norms expressed in schools have been shown to play a powerful role in shaping behaviors and health outcomes (58,59), and integrating WASH interventions with normative behavior change has shifted norms and behaviors in other contexts (60). It is also important to acknowledge that most WASH research is led by academics in engineering and medicine, often with a bias toward cognitive theories of behavior change. Collaboration with colleagues in the social sciences will be crucial to overcome gaps in domain specific expertise and to expand the hypotheses we form and test beyond current disciplinary boundaries. Future research should seek to understand if normative behavior change strategies can play a role in promoting healthy WASH practices in schools.

## ACKNOWLEDGEMENTS

Funding for this project was provided by World Vision and doctoral funding for Gracie Hornsby was provided by Knight-Hennessy Scholars. We thank Jordan Smoke, Kristie Urich, Ben Tidwell, and Peter Hynes of World Vision for their continual support. We would also like to thank all of the enumerators who worked on this project and made this work possible. Thank you to all of the students, school staff, families, and government officials who participated in this study. Thank you to Swati Agnihotri for your leadership at World Vision-India, as well as Sarita Dubey, Anurag Mishra, Pratima Verma, Amit Singh, and Govind Singh. Thank you to the many Stanford students who contributed to this project over the years: Hadassah Betapudi, Rohan Chandran, Sarika Lansberg, and Sam Wind. A special thanks to Jenelle Van Eynde and Carine Sauquet for logistical support that kept this project on track.

## FOOTNOTES

### Contributors

JD and GD are the co-principal investigators of the study and responsible for funding acquisition, supervision, and the overall content as guarantors. JD, GD, JW, and CP conceptualised the research. All co-authors contributed to drafting measurement instruments, and DB, PS, RS, AB, and GH piloted them. GH drafted the pre-analyses plan and conducted all analyses, coding, and visualisations with support from PS and PG. All authors contributed to the interpretation of results and discussion. GH drafted the original manuscript in collaboration with JD and GD. All authors contributed to the critical review and editing of the manuscript. All authors reviewed and approved the final version of the manuscript. The authors alone are responsible for the views expressed in this article, which do not necessarily reflect the views, decisions or policies of their affiliated organisations.

### Funding

This work was supported by World Vision USA

### Competing interests

All authors have completed the Unified Competing Interest form (available on request from the corresponding author) and declare: no support from any organisation for the submitted work; no financial relationships with any organisations that might have an interest in the submitted work in the previous three years, no other relationships or activities that could appear to have influenced the submitted work.

### Data availability statement

Data for this project have not yet been released publicly, but can be made available upon reasonable request.

### Structured reflexivity statement

#### Study Conceptualization

This study was motivated by the policy and programmatic priorities of World Vision-India, who worked in close partnership with the state government of Uttar Pradesh. Government officials were engaged and updated throughout the research process. Local researchers played a leadership role in designing formative work and qualitative midline and endline assessments.

#### Research Management

Local research teams were supported for travel to the field, all billable hours of work, as well as for analysis and publishing activities.

#### Data Acquisition and Analysis

Research staff who oversaw data collection efforts are included as co-authors on this paper. Both organizations in this research partnership have full access to the data that was collected. Local researchers indicated their primary interest was in analysis of the qualitative data collected. They played a leadership role in selecting the methods for thematic analysis, as well as in preparing reports and manuscripts presenting qualitative research findings.

#### Data Interpretation

Research partners met multiple times to present and review data throughout analysis. Both teams gave feedback and informed the interpretation of the analysis and conclusions.

#### Drafting and revising for intellectual content

Research partners indicated particular interest in leading on analysis and publication of the qualitative data collected during formative work and midline and endline assessments, given their expertise in qualitative research. Research partners received regular feedback on their writing from co-authors to refine drafts. Research products will be adapted for local needs into short research briefs currently in preparation for distribution to schools and local government. Local researchers led a workshop with district-and state-level stakeholders to discuss findings and recommendations.

#### Authorship

Research partners, given their expertise in qualitative research, serve as the first and last authors on publication of the qualitative formative work (in press, cited in this paper). They are also identified as co-authors on this work. This authorship arrangement was discussed prior to the onset of data analysis, and a publication plan was written collaboratively by both teams.

Early career researchers are included as both first and co-authors on every publication from this trial. Authorship has been assigned based on ICMJE standards for co-authors. After following these standards, the gender balance of our authorship team reflects the labor that was contributed to this trial.

#### Training

Our research partners are highly trained professional researchers who hold MS, MPH, MD, and PhD degrees. Researchers were offered the opportunity to expand their skillsets during data analysis and writing, as their workloads and interests allowed.

#### Infrastructure

India already has a strong local infrastructure for research and critical inquiry. Through this project, researchers gained additional skills that will equip them for future investigations.

#### Governance

All study staff followed local guidelines for preventing the spread of COVID-19 at the beginning of the study period when local transmission rates were comparatively higher. The highest precautions have been taken to anonymize data so schools cannot be identified from this research, particularly in research briefs distributed to local stakeholders.

## Notes

### Competing Interest Statement

The authors have declared no competing interest.

### Clinical Trial

RIDIE-STUDY-ID-662bed52ae1e7

### Author Declarations

IRB of Stanford University gave ethical approval for this work (protocol 69912) IRB of Sigma Research and Consulting gave ethical approval for this work(protocol 10046/IRB/22-23)

