## Supplementary for "The individual and combined impacts of school-based water, sanitation, and hygiene (WASH) education and infrastructure operation and maintenance (O&M): A factorial cluster-randomized controlled trial in Uttar Pradesh, India"

**Section S1. Supplementary Figures and Tables**

***Table S1.*** *Definitions for intermediate outcomes*

| **Intermediate Outcome** | **Definition** |
| --- | --- |
| **Student Knowledge** | |
| Student knowledge of handwashing before eating | Student reports “before eating” as an important time to wash hands in response to an open-ended question |
| Student knowledge of handwashing after using the toilet | Student reports “after using the toilet” as an important time to wash hands in response to an open-ended question |
| Student knowledge of germs | Student reports information about germs that is accurate AND relevant to WASH/health/science in response to an open-ended question. Accurate is defined as scientifically factual information about germs. Relevant is defined as information that pertains to healthy WASH behaviors, WASH infrastructure, health, or curriculum learning outcomes |
| **Infrastructure & Consumables** | |
| Handwashing water availability | Percent of schools where unannounced observers found at least one functional source of water for handwashing |
| Soap availability | Percent of schools where unannounced observers found soap visibly available to students |
| Toilet accessibility | Percent of schools where unannounced observers found at least one of the toilet stalls that was randomly selected for observation was unlocked |
| Toilet functionality | Percent of schools where unannounced observers found at least one of the toilet stalls that was randomly selected for observation was able to flush/drain properly when tested by the observer |
| Anal cleansing water availability | Percent of schools where unannounced observers found at least one of the toilet stalls that was randomly selected for observation had adequate water available for anal cleansing |
| Toilet cleanliness | Among toilets randomly chosen for observation, the average toilet cleanliness score. The toilet cleanliness score is a composite indicator scored 0-8 where each of four components are scored 0-2 (0 = A lot, 1 = Some, 2 = Very little/None) and weighted equally. The four components are presence of (1) visible feces, (2) mud/dirt/trash, (3) pest and flies, and (4) unpleasant odor. |

***Table S2.*** *Checks for selective attrition by based on block, student gender, student grade, soap access at home, and toilet access at home. We detected evidence of selective attrition by block, gender, and grade. These variables were modeled as fixed effects to mitigate the effect of selective attrition on model estimates.*

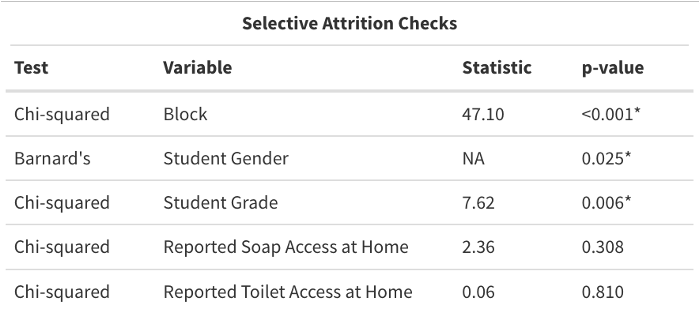

***Table S3.*** *Baseline balance checks were performed to detect imbalance between treatment arms on key variables. A significant effect for observed toilet presence was detected between study arms. All schools had structures containing toilets at baseline, however 1-3 schools in each treatment arm were performing temporary construction on their toilets at baseline, thus they were unavailable to students or observers. More details reported in S5.*

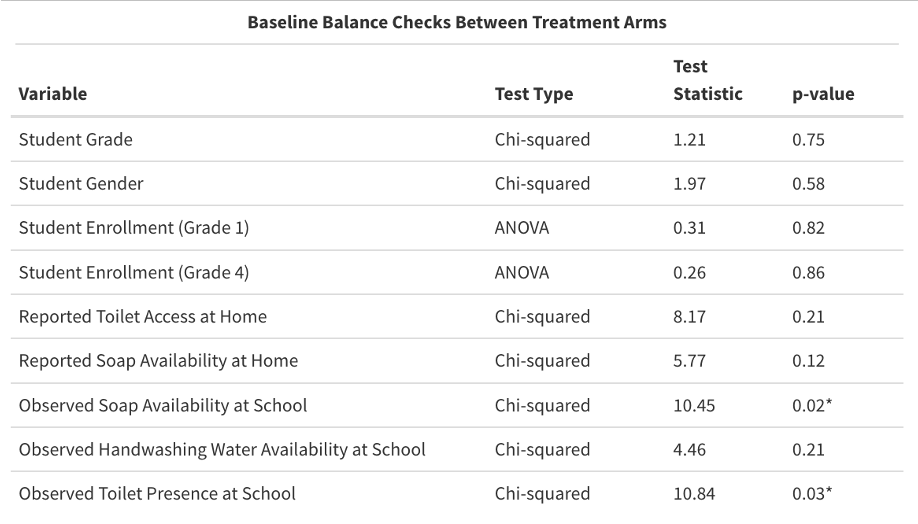

***Table S4.*** *Summary of analytical approach*

| **Outcome** | **Model** | **Fixed Effects** | **Random Effects** |
| --- | --- | --- | --- |
| **Student Knowledge** | | | |
| Student knowledge of handwashing before eating | Generalized linear mixed model (glmer) | Treatment group, time, student gender, block ID | Cluster ID |
| Student knowledge of handwashing after using the toilet |  | Treatment group, time, student gender, block ID | School ID, Cluster ID |
| Student knowledge of germs |  | Treatment group, time, student gender, student grade, block ID | School ID, Cluster ID |
| **Enabling Environment** | | | |
| Handwashing water availability | Insufficient variability; descriptive statistics only | NA | NA |
| Soap availability |  | NA | NA |
| Toilet accessibility | Generalized estimating equation (GEE) | Treatment group, time | School ID |
| Toilet functionality |  | Treatment group, time, block ID | School ID |
| Anal cleansing water availability |  | Treatment group, time, block ID | School ID |
| Toilet cleanliness | Generalized additive models for location, scale, and shape (GAMLSS) | Treatment group, time, block ID | School ID |
| **Student Behavior** | | | |
| Observed rate of student handwashing | Generalized Linear Mixed Models using Template Model Builder (glmmTMB) | Treatment group, time, block ID | School ID |
| Observed rate of student toilet use |  | Treatment group, time | School ID |

***Table S5.*** *Characteristics of schools at baseline measurement. Student-level variables are aggregated within each school, then reported as treatment-level averages.*

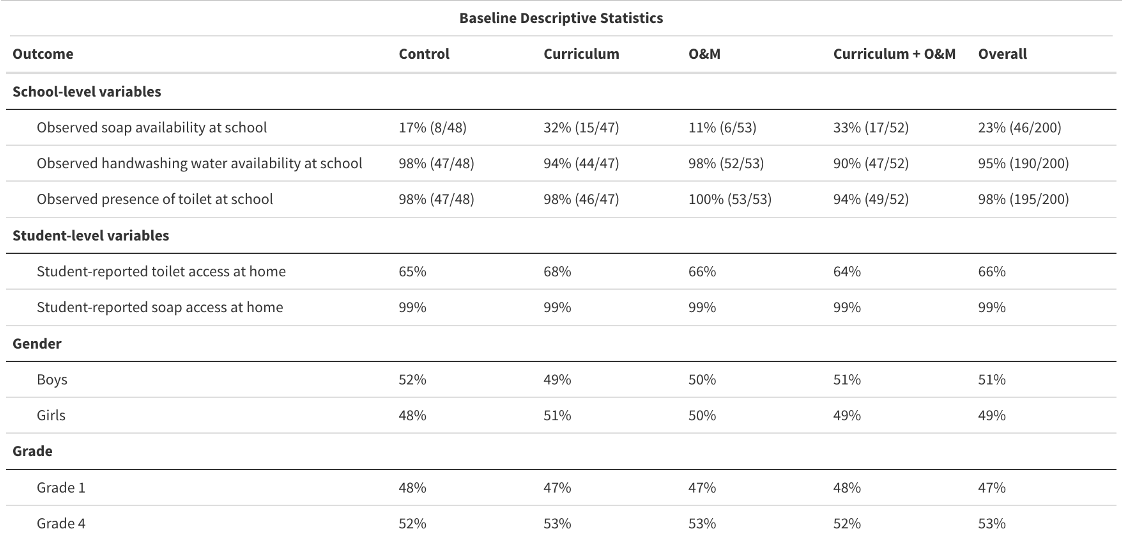

***Figure S1.*** *Soap availability as an illustrative example of the observed-induced bias in infrastructure outcomes at baseline, midline, and endline measurements. These measurements were taken on the same day as student and teacher surveys, meaning these announcements were not unannounced since schools were informed of what day student surveying would occur.*

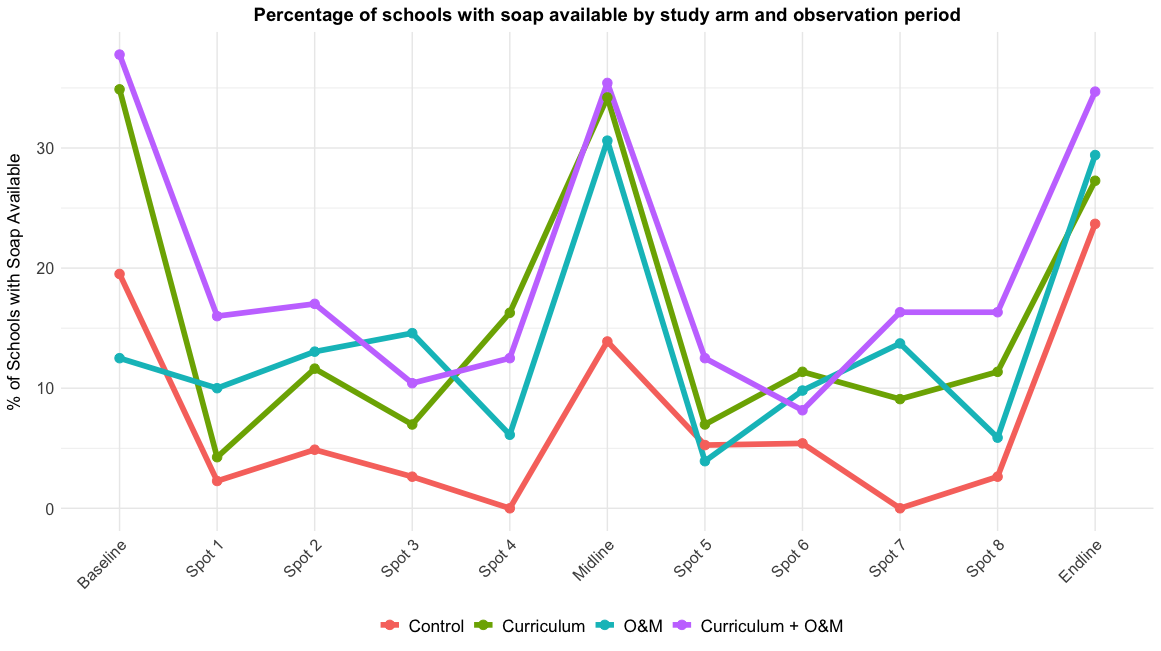

***Figure S2.*** *Forest plot of model results for student knowledge of handwashing before eating. Results show that girls started with significantly lower knowledge at baseline, but improved at a similar rate as boys over time.*

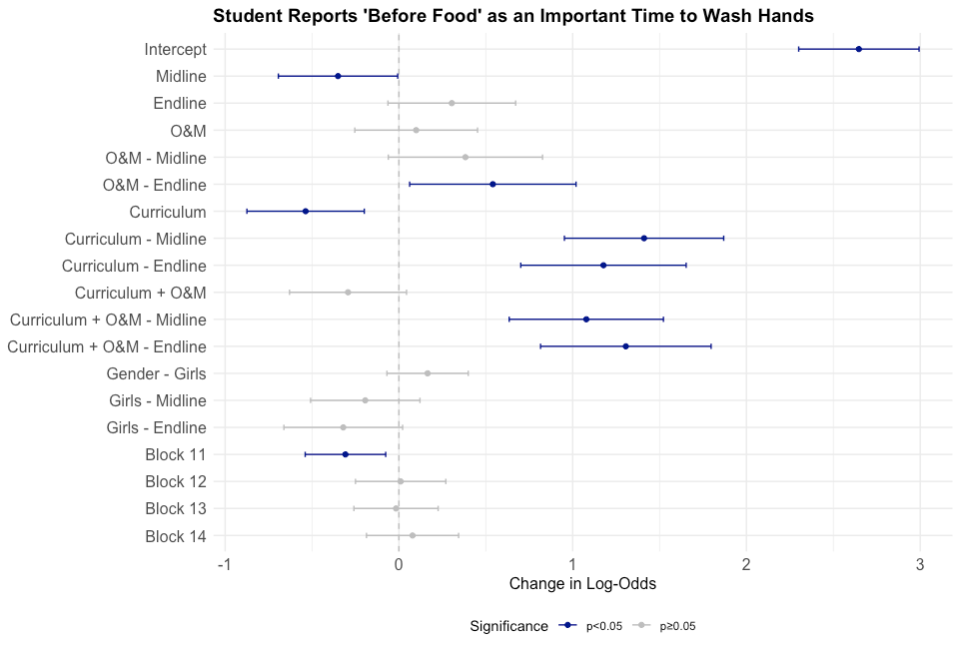

***Figure S3.*** *Forest plot of model results for student knowledge of handwashing after using the toilet. Results show that girls and boys started with comparable knowledge at baseline and improved at a similar rate over time.*

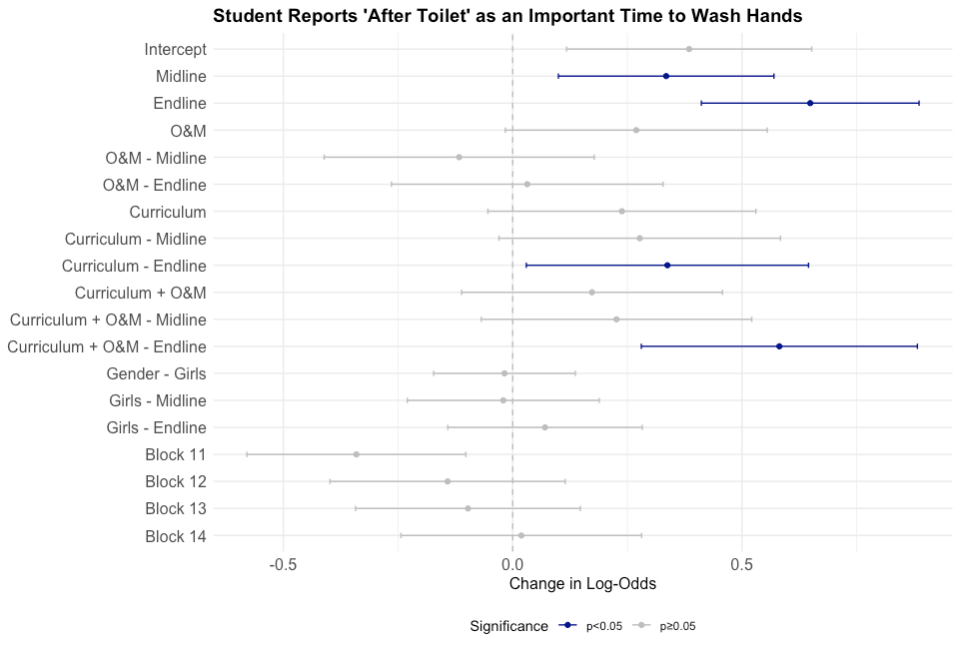

***Figure S4.*** *Forest plot of model results for student knowledge of germs in response to an open-ended question. Results show that girls started with significantly lower knowledge at baseline, but improved at a similar rate as boys over time. Results also show that grade one students started with significantly worse knowledge at baseline. There is a significant grade-by-time interaction at midline and endline indicating grade one students had significantly larger change in long-odds than grade four. However, this change in log-odds equates to a similar change in probability in each group due to the non-linear relationship between log-odds and probability.*

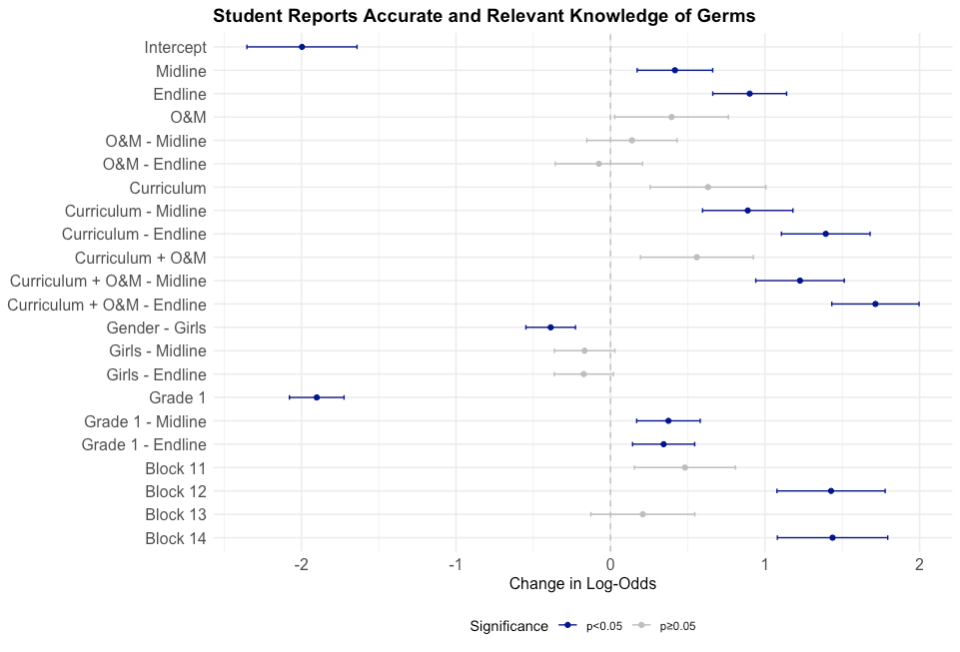

***Table S6.*** *Data quality check to ensure students were not coached to give certain interview answers by school staff or parents. Coaching was minimal and not differential across treatment arms.*

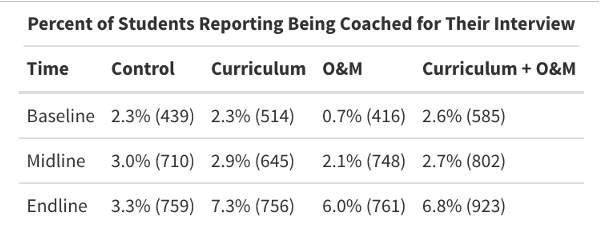

***Figure S5.*** *Time-varying model results in units of predicted probability for four toilet-related intermediate outcomes: toilet accessibility (unlocked), toilet cleanliness score, toilet functionality (able to drain properly), and availability of water for anal cleansing in the toilet stall. Toilet cleanliness is reported in terms of the toilet cleanliness score (0-8) instead of predicted probability. “BL” refers to baseline measurement and all numbered time points on the x-axis refer to unannounced infrastructure observations between rounds of surveying. Midline measurements were taken between time points 4 and 5, and endline measurements were taken after time point 8.*

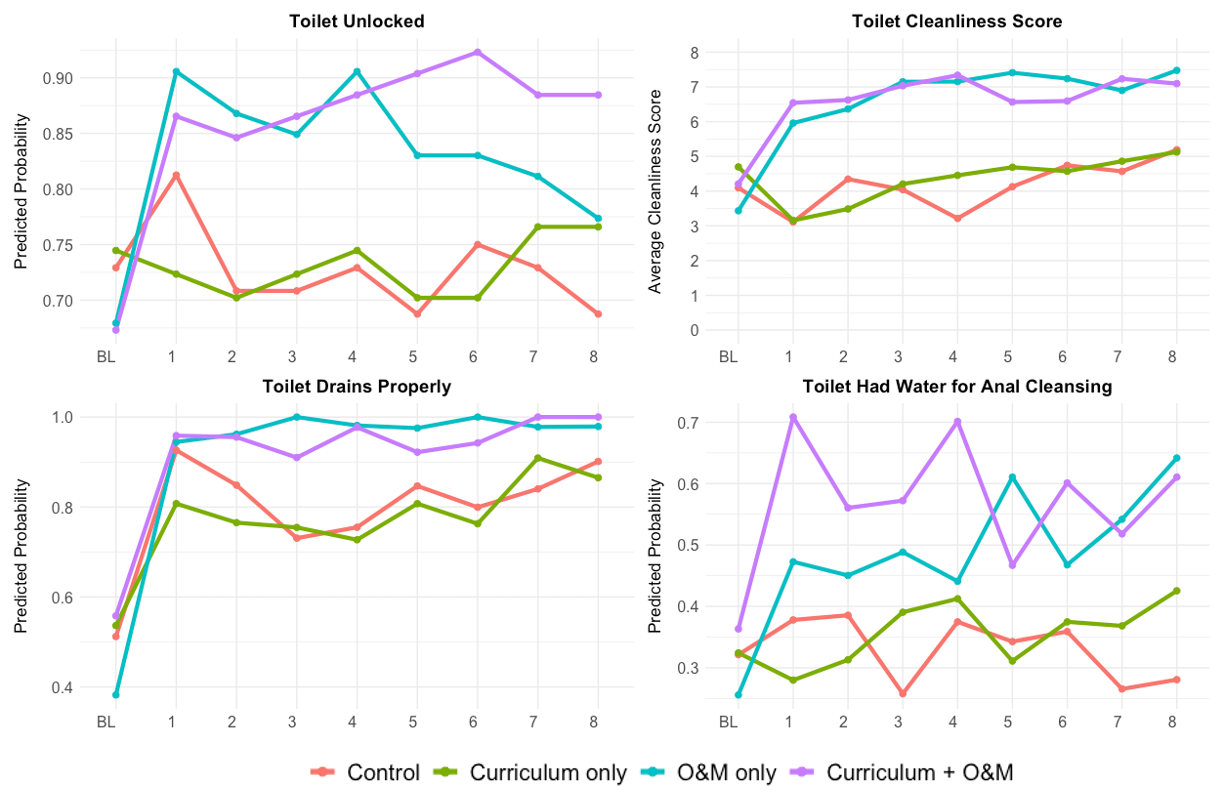

**Section S2. Additional intervention details**

We undertook formative research to inform the O&M intervention design, including semi-structured interviews with school staff (n=34), government cleaners (n=9), parent members of school management committees (n=15), and local government officials (n=12). Each cleaner worked for approximately two hours per day, scheduled by mutual agreement with the school to which they were assigned. All cleaners were contracted by a local, private third-party agency which trained and monitored the performance of cleaners, handled any complaints from schools, and restocked cleaning materials like brushes and bleach, and was subject to financial penalties for poor- or non-performance.

**Section S3. Power calculations**

This study is powered to detect a minimum detectable effect (MDE) size of 0.169 for difference-in-differences (DiD) analyses, related to students' knowledge, and 0.071 for repeated measures ANOVA, associated with observational data on infrastructure and student behavior. We designed for a significance level of 0.05 with a targeted power of 0.80 (beta=0.2). Variability in effect size is expected to stem from intracluster correlation (assumed ICC ≤ 0.05), baseline differences, grade levels, and gender. We accounted for ICC in our power calculations, and we expect baseline variability to be approximately randomly distributed due to random selection of schools and random treatment allocation. We also drew a sample with even grade-level and gender representation to allow subsetted analyses among these groups.

With one site and 200 schools, the study anticipates explaining a moderate share (0.3-0.6) of variance with the covariates identified in this plan. Searching prior experimental literature on the impact of WASH improvements on behavior and infrastructure outcomes, we find low R-squared values generally ranging between 0.1-0.28. We expect to exceed the predictive ability of previous trials because of the higher degree of control in this trial (normalized baseline infrastructure conditions, cluster randomization) and the quality of the data collected.

Sensitivity analyses reveal the MDE for DiD analyses is somewhat responsive to an increase in the expected ICC but robust to attrition variations. If the ICC is higher than estimated (0.05), we can detect an effect size of 0.218 with an (high) ICC of 0.10 and an effect size of 0.293 with an (very high) ICC of 0.2. For repeated measures ANOVA, the MDE is again somewhat sensitive to changes in the ICC. An (high) ICC of 0.10 and (very high) ICC of 0.20 could increase the MDE to 0.082 and 0.110, respectively. Repeated measures ANOVA analysis are once again robust to attrition and variations in the average sample size per school.

**Section S4. Sampling Frame**

Leveraging government data on the more than 6000 government schools in the study district, we identified the roughly one-third that served primary students (grades one to five) and were recorded in India’s national Unified District Information System for Education (UDISE) database as having functional, gender-segregated sanitation facilities and an improved water source (Figure 2). We selected five blocks in the district with probability of selection proportional to the number of schools in a given block. Within each block, we drew a simple random sample of 49 schools to form a preliminary study sample of 245 schools. We visited each school to verify the presence and functionality of WASH facilities, as well as school enrollments. We excluded schools that lacked an improved water source and/or gender segregated sanitation facilities (n=23); had an enrollment of fewer than 71 students (n=19); required WASH infrastructure repairs that exceeded WV’s budget (n=13), or were located in a very remote area (n=1). To achieve the minimum required sample size of 200, we added eleven non-randomly selected schools that met the study criteria and were identified by block education officers.

Twenty-seven (14%) schools received handpump repairs; 129 (64%) received storage tanks to enable construction of handwashing stations; 60 (30%) had their toilets unclogged; and 94 (47%) received replacement urinal pots and/or squatting pans in their toilets. Repairs and upgrades were completed at schools in early 2020, after which the study was paused for over two years due to the COVID-19 pandemic. Given this long delay, we re-evaluated the presence of toilets and an adequate water source on school premises in baseline balance checks.

**Section S5. Treatment assignment**

Within each block, we created four clusters of 8-13 schools each. Each cluster included schools that were in close proximity, not separated by major geographical barriers like roads or rivers when possible, and not all in block centers or the most isolated areas of the block.

GH performed treatment allocation and then was blinded to treatment identities by partners at Oxford Policy Management. Treatment assignments were stored in a password protected spreadsheet and were not accessed after the initial allocation until all pre-specified analyses were completed.

**Section S6. Random student selection**

Prior to student surveys, we distributed parental consent forms to all schools for local teachers to explain and share with parents. Students whose parents returned a signed consent form within the designated time frame were eligible for selection into the study. Forms were randomly drawn to create an ordered list of student names by grade. At each school, the first 20 grade one and 20 grade four students on the list who were present on the day of surveying and assented to participate comprised our baseline sample. To minimize the effect of attrition and optimize longitudinal sampling, we replaced students lost to follow-up with any students who previously missed a round of surveying but were again available for surveying. If no such student was available, we randomly selected another student from the same school, grade, and gender (Figure 2).

**Section S7. Extended data analysis details**

To mitigate potential impacts of selective attrition by block that we identified during balance checks (Table S2), we modeled block as a fixed effect rather than a random effect and dropped the covariate if block effects were not significant or model fit was not improved, as was specified in our pre-analysis plan. We also modeled gender and grade as fixed effects for student-level outcomes and retained them in the model when they improved model fit. We omitted missing data points from analysis, but retained the remaining data for a given student or school. All models leveraged in this analysis use maximum likelihood estimation (MLE) to estimate across students or schools with incomplete data, except for generalized estimating equation (GEE) models which use quasi-likelihood estimation.

We modeled knowledge outcomes using a generalized linear mixed effects model with a random effect for school ID and fixed effects for treatment condition, time, and block. We modeled infrastructure outcomes, except for toilet cleanliness, using a mixed-effects GEE model with a random effect for school ID and fixed effects for treatment condition, time, and block. Toilet cleanliness was modeled using a generalized additive model for locations, scale, and shape (GAMLSS) because the data distribution violated assumptions for GEE. Lastly, we modeled behavior outcomes using generalized linear mixed models using Template Model Builder (glmmTMB) to handle overdispersion in the data. Models contained a random effect for cluster ID and a fixed effect for block when it improved model fit.

**Section S8. Statistical adjustment of baseline values**

Significant observer-induced bias was identified in some of our infrastructure outcomes (soap availability, toilets unlocked, toilet cleanliness) at baseline, midline, and endline. For example, soap availability was on average 19 percentage points higher at scheduled baseline, midline, and endline measurements (28%) than at unannounced observations between major rounds of surveying (9%) (Figure S3).

Baseline values for two intermediate outcomes, toilets unlocked and toilet cleanliness, displayed this clear bias from schools responding to observation at baseline measurements. Schools responded to observation by unlocking toilets more often and cleaning toilets prior to our team’s arrival. Toilets quickly became locked and dirtier in the Control arm in the following measurements taken at unannounced spot checks, indicating baseline measurements were likely not reflective of schools’ actual pre-intervention behavior. Pre-adjustment baseline values are shown in Figures S6 and S7.

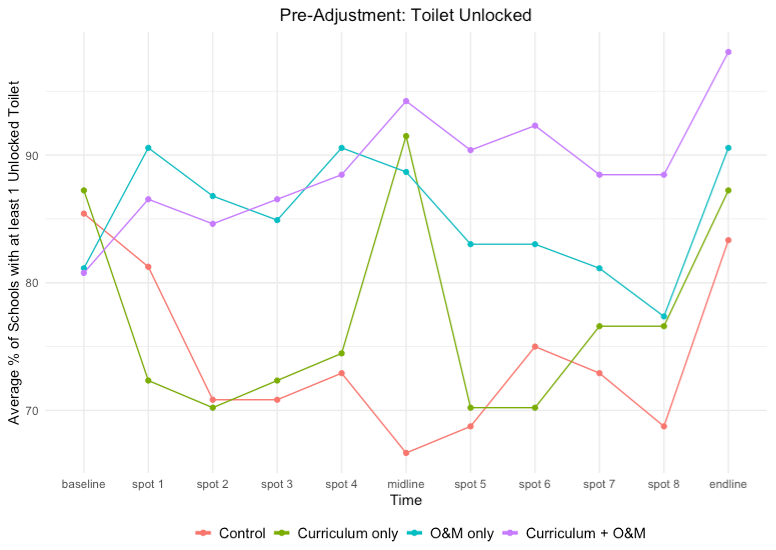

***Figure S6.*** *Data for toilet accessibility prior to baseline adjustment. Data are expressed as treatment-level averages, not modeled output.*

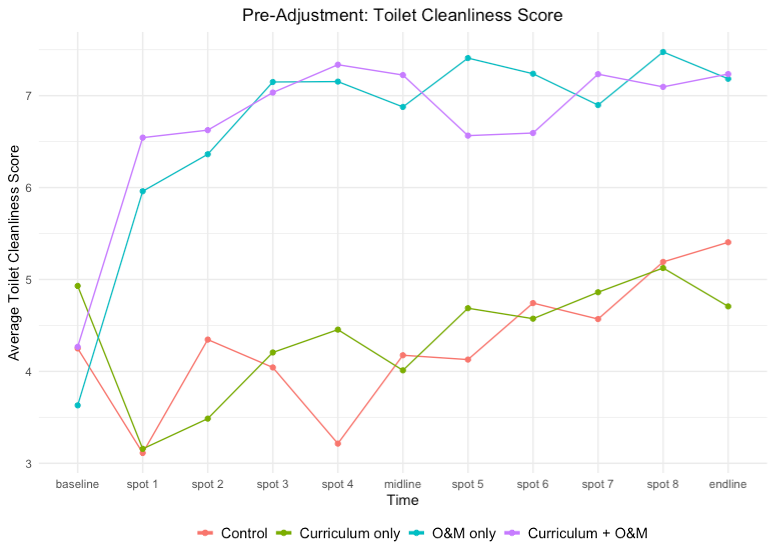

***Figure S7.*** *Data for toilet cleanliness prior to baseline adjustment. Data are expressed as treatment-level averages, not modeled output.*

Baseline is an influential data point for difference-in-difference calculations since change is in reference to the baseline data point for each arm. Thus, bias in baseline data points can mask intervention effects and lead to inaccurate conclusions. To adjust for the observed bias, we calculated the mean and standard deviation (SD) of the Control group at baseline as well as the mean value of unannounced spot checks 1-8. We subtracted the mean value from unannounced spot checks from the baseline mean. We used this difference to represent the estimated baseline bias. To perform this adjustment, we had to assume bias was evenly distributed across schools, regardless of treatment.

To adjust baseline values, we calculated the baseline mean and SD of each arm at baseline. We subtracted the estimated baseline bias from each arms’ baseline mean. To allow the data distribution to vary across treatment arms and reflect the pre-adjustment distribution, we used R to randomly re-assign binary baseline values to each school to achieve the correct adjusted mean and original SD in each study arm. Midline and endline values were also omitted from all infrastructure-related outcomes to limit bias in the dataset. Post-adjustment baseline values are shown in Figures S8 and S9.

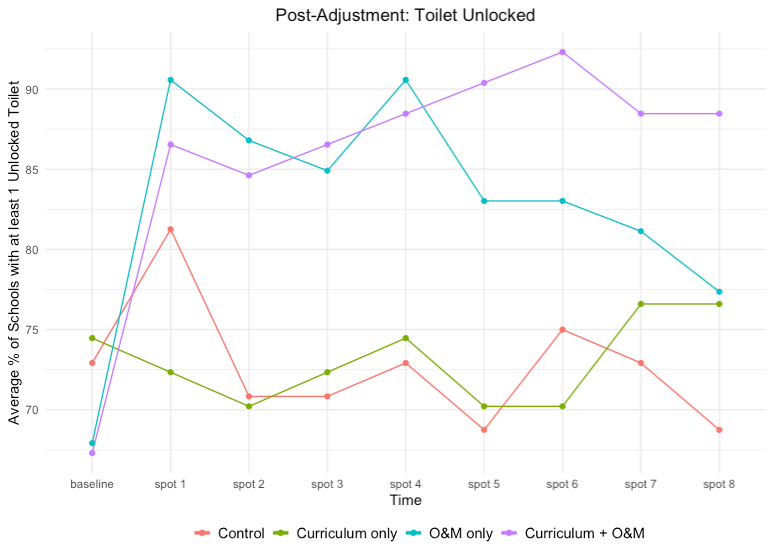

***Figure S8.*** *Data for toilet accessibility after the baseline adjustment. Data are expressed as treatment-level averages, not modeled output.*

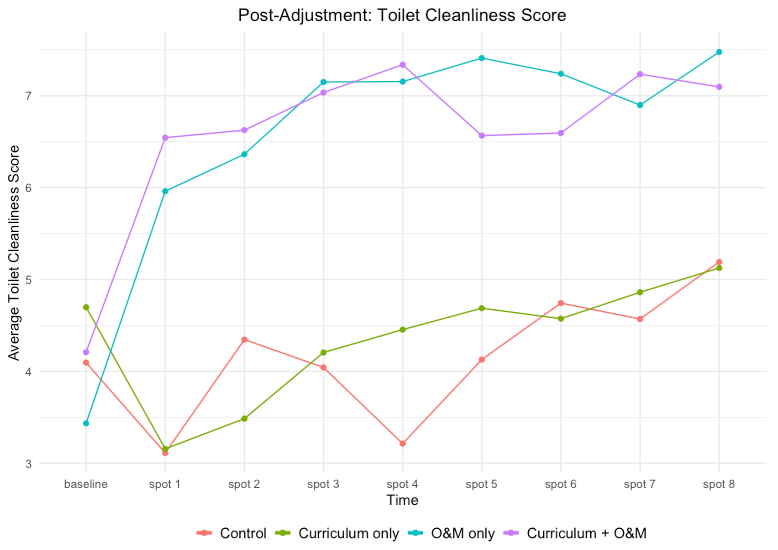

***Figure S9.*** *Data for toilet cleanliness after the baseline adjustment. Data are expressed as treatment-level averages, not modeled output.*

**Section S9. Toilet cleanliness score definition**

The toilet cleanliness score is a composite indicator of four observed cleanliness metrics: presence of visible urine/feces in the toilet stall outside of the toilet pan/bowl, presence of mud/dirt/trash in the toilet stall, presence of flies or other pests in the toilet stall, presence of unpleasant odors in the toilet. Each of the four metrics were scored “a lot” (0), “some” (1), “very little/none” (2) so that a higher score indicates a cleaner toilet.

Observers were uniformly trained on the scoring system through pictures and had a chance to ask questions and refine their understanding during pilot testing and training.

**Section S10. Sensitivity analysis and quality check results**

We performed a sensitivity analysis after removing the 11 government-selected schools from the sample and produced similar results on all aforementioned outcomes. We also re-analyzed observational behavior data after stratifying by morning versus afternoon time periods and observed no notable differences. We included questions in the student surveys that assessed whether students had been coached to give certain answers or prepared for their interview in any way. Student reports of preparation in advance of surveys by school staff or parents were minimal (0.7%-7.3% of students) and approximately evenly distributed across study arms (Table S6).
